# Unpacking sources of transmission in HIV prevention trials with deep-sequence pathogen data – BCPP/ Ya Tsie study

**DOI:** 10.1101/2024.08.30.24312845

**Authors:** Lerato E. Magosi, Eric Tchetgen Tchetgen, Vlad Novitsky, Molly Pretorius Holme, Janet Moore, Pam Bachanas, Refeletswe Lebelonyane, Christophe Fraser, Sikhulile Moyo, Kathleen E. Hurwitz, Tendani Gaolathe, Ravi Goyal, Joseph Makhema, Shahin Lockman, Max Essex, Victor De Gruttola, Marc Lipsitch

## Abstract

To develop effective HIV prevention strategies that can guide public health policy it is important to identify the main sources of infection in HIV prevention studies. Accordingly, we devised a statistical approach that leverages deep- (or next generation) sequenced pathogen data to estimate the relative contribution of different sources of infection in community-randomized trials of infectious disease prevention. We applied this approach to the Botswana Combination Prevention Project (BCPP) and estimated that 90% [95% Confidence Interval (CI): 81 – 93] of new infections that occurred in individuals in communities that received combination prevention (including universal HIV test-and-treat) originated from individuals residing in communities outside of the trial area. We estimate that the relative impact of the intervention was greater in rural geographically isolated communities with limited opportunity for imported infections compared to communities neighboring major urban centers. Treating people with HIV limits the spread of infection to uninfected individuals; accordingly, counterfactual modeling scenarios estimated that a nationwide application of the intervention could have reduced transmissions to recipients in trial communities by 59% [3 – 87], much higher than the observed 30% reduction. Our results suggest that the impact of the BCPP trial intervention was substantially limited by sources of transmission outside the trial area, and that the impact of the intervention could be considerably larger if applied nationally. We recommend that studies of infectious disease prevention consider the impact of sources of transmission beyond the reach of the intervention when designing and evaluating interventions to inform public health programs.

## Introduction

Why did the landmark community-randomized universal HIV test-and-treat trials in sub-Saharan Africa - BCPP/ Ya Tsie [1], HPTN 071/ PopART [2], SEARCH [3] and ANRS 12249/TasP [4] - show variable reductions in the occurrence of new HIV infections in trial communities that received the intervention compared to control communities (0% – 30%) despite substantial gains in viral suppression [5]? This is one of the most important questions in the HIV policy world today because HIV ‘test-and-treat’ was thought to hold great potential to bring the HIV epidemic under control in the absence of a successful vaccine or functional cure. Some of the variation in the incidence reductions observed is thought to be due to a change in national HIV treatment guidelines to universal treatment part-way through the trials effectively reducing the difference between intervention and control communities. Another complementary hypothesis is that HIV transmissions to residents of intervention communities from individuals in non-intervention communities in the trial (control communities) and from communities not taking part in the trial (non-trial communities) limited the size of effect observed in the trials, but it is unknown to what degree. A large dilution of the intervention effect in the trials by transmission from non-intervention communities could suggest a larger impact of the intervention than originally envisaged [6-8].

We test this hypothesis in one of the four trials, the Botswana Combination Prevention Project. Specifically, we developed a statistical modeling approach that uses directed sexual contacts inferred from deep-sequenced HIV virus to estimate the relative extent to which transmissions in trial communities occurred from individuals in the same community; different communities in the same trial arm; different communities in the opposite trial arm; and non-trial communities. In addition, to estimate what the impact of a nationwide intervention would have been on recipients in trial communities, we apply our statistical approach in “counterfactual” modeling scenarios that estimate the relative contribution of the different sources of infection mentioned above in the presence and absence of the intervention. Note that in a nationwide intervention all communities nationally would receive combination prevention (including universal HIV test-and-treat).

## Materials and Methods

### BCPP Study Description and Data

#### BCPP study description

The Botswana Combination Prevention Project (BCPP, also known as the Ya Tsie trial) was a pair-matched community-randomized trial to evaluate the effect of universal HIV testing and treatment on HIV incidence reduction. The trial was conducted in 30 rural and peri-urban communities across Botswana from 2013-2018 [1]. Trial participants were adults aged 16-64 years and the average population size eligible to participate in each trial community was 3,820 people. Communities were matched into 15 pairs based on three criteria: geographical proximity to major urban areas (Gaborone city, Palapye and Francistown city), population size and age structure, and access to health services; then within each pair, communities were randomized into the intervention and control arms of the trial. The 15 intervention communities in the trial received expanded access to universal HIV testing (with attempt to test all willing adult residents who did not have documented positive HIV status), strengthened linkage-to-care for early treatment, and expanded treatment availability. After a period of community sensitization through door-to-door canvassing, community leadership engagement and public loudspeaker announcements, mobile and home-based HIV testing campaigns were conducted within each intervention community over approximately two consecutive months [9]. Routine testing in intervention community health facilities was reinforced to diagnose all people with HIV and avail them early treatment. An additional effort was made to offer HIV testing to men and youth where they work and socialize, for example: at bars and community football (soccer) matches. To strengthen linkage-to-care, people with HIV who were not on treatment were assisted to schedule an appointment at a local clinic, provided text alerts prior to the appointment and followed-up to re-schedule in the case of a missed appointment. Access to services for safe male circumcision and prevention of mother-to-child transmission was also expanded in intervention communities. By comparison, control communities received the standard-of-care, which before 2016 meant that people with HIV qualified to start antiretroviral treatment when their CD4 cell count was below 350 cells per microliter. Beginning June 2016, the national HIV treatment policy was changed to universal treatment meaning that immediate antiretroviral treatment was now available in both arms of the BCPP trial. To evaluate HIV incidence reduction, an HIV incidence follow-up cohort was established through a baseline household survey of a random sample of 20% of households in each trial community. Annual household surveys with retesting for HIV (in persons who were HIV-negative) were then conducted in the same 20% household sample in all 30 communities during the trial. The BCPP trial comprised 7.6% (175,664) of the national population and showed a 30% reduction in the occurrence of new HIV infections in intervention communities compared to control communities over an average of 29 months [1]. In addition, the BCPP trial conducted an end-of-study survey of 100% of households in 3 intervention communities and 3 control communities to assess progress on the 90-90-90 UNAIDS targets. Trial participants with HIV were invited to provide a sample for viral phylogenetic analysis. This included all people with HIV from (1) the baseline household survey, (2) annual household surveys, (3) end-of-study survey, as well as (4) all people with HIV (but not yet on ART) who were referred for treatment during community-wide testing and counseling campaigns, (5) all people with HIV that later presented at health care facilities in intervention communities and (6) all people with HIV who were already receiving HIV care at health facilities in intervention communities.

#### Deep-sequence phylogenetics data

Near full-length genome sequences were obtained using predominantly proviral DNA (as the majority of study participants were virally suppressed on ART) or RNA. The HIV-1 viral consensus whole genomes of individuals that met minimum criteria for inclusion in phylogenetic analyses were ones that had fewer than 30% of bases missing beyond the first 1,000 nucleotides [10]. To efficiently use computational resources, viral consensus whole genomes were used to identify groups (or clusters) of trial participants with genetically similar HIV-1 infections as a filtering step to exclude distantly related sequences from deep-sequence phylogenetic analysis [10]. A detailed description of the deep-sequence phylogenetic analysis is published in [10]. Briefly, we performed ancestral host-state reconstruction with the phyloscanner software [11, 12] to identify pairs of trial participants with genetically similar HIV-1 infections and the probable direction of transmission between them (female-to-male or male-to-female). For brevity, we refer to the directed opposite-sex transmission pairs as source-recipient pairs. For each identified transmission pair there was accompanying metadata on the names of the communities in which the source and recipient partners reside and the randomization-condition to which their communities were assigned.

#### Pairwise drive distance data

Pairwise drive distances between ordered pairs of 488 communities in the 2011 Botswana population and housing census were successfully sourced from the google distance matrix application programming interface (API) with the mapsapi package v0.5.0 in R v4.1.1 [13]. The 488 census communities included all 30 communities in the BCPP trial. Therefore, of the possible 488 × 488 ordered community pairs between the 488 census communities we sourced 488 × 30 ordered community pairs that had any of the 488 census communities as a source (origin) community and any of the 30 trial communities as a recipient (destination) community.

#### Population-size and HIV prevalence estimates

Population-size estimates of 488 communities in the 2011 Botswana population and housing census were sourced from [14], and district-level HIV prevalence estimates were obtained from the 2013 Botswana AIDS Impact Survey (BAIS 2013) [15].

### Estimating Transmissions to Recipients in BCPP Trial Communities

To estimate transmissions that occurred to recipients in trial communities from different sources of infection nationally, we first inferred directed (opposite-sex) transmission events between the 30 BCPP trial communities using deep-sequence phylogenetics. Then we statistically modeled the risk of transmission between trial communities using a negative-binomial regression framework; with the inferred transmission events as the response variable and the following variables as predictors: (1) the pairwise drive distance separating the source and recipient communities, (2) whether the source community was randomized to receive the intervention, and (3) whether the source community and the recipient community were the same (within-community transmission) or different (between-community transmission). After that we used the pairwise drive distances between the 488 communities in the 2011 Botswana population and housing census as input to the model of the risk of transmission between trial communities to predict the risk of transmission (expected probability of viral genetic-linkage had the cases been sequenced) between communities nationally. Finally, to estimate the number of transmissions into trial communities from all communities nationally, estimates of the risk of transmission to recipients in trial communities from communities nationally were combined with population-size estimates from the 2011 Botswana population and housing census [14] and district-level HIV prevalence estimates from the 2013 Botswana AIDS Impact Survey (BAIS 2013) [15]. The supplementary appendix provides a detailed account of the statistical approach used to estimate the relative contribution of different sources of infection in: 1) the same community, 2) different communities in the same trial arm, 3) different communities in the opposite trial arm and 4) in communities outside the trial area.

## Results

Of the 5,114 trial participants who consented to a blood draw for viral genotyping and whose HIV viral whole genomes were successfully deep-sequenced [1, 10], 3,832 met inclusion criteria for phylogenetic analysis, and from those, we identified 82 directed opposite-sex transmission pairs between ordered pairs of the 30 communities in the BCPP trial (Supplementary Figure 1) [10]. Of the 82 source-recipient pairs, 51 (21 female-to-male, 30 male-to-female) were identified between HIV viral genomes sampled during the baseline period of the trial compared to 31 (16 female-to-male, 15 male-to-female) where the recipient’s genome was sampled post-baseline. We defined the post-baseline period as at least one year after baseline household survey activities had concluded in a community such that the intervention could have taken effect.

### Relationship between the drive distance separating pairs of communities and the risk of transmission between them

We first demonstrate a relationship between the drive distance separating communities in the BCPP trial and the risk of HIV-1 transmission between them (Table 1 and Figure 1). We define the risk of transmission as the expected probability of viral-linkage between deep-sequenced HIV viruses of individuals with HIV randomly sampled from their respective communities. Figure 1 and Table 1 show that the risk of transmission decreases as the drive distance separating community pairs increases, specifically by 27% [95% Confidence Interval (CI): 3 - 45] per 100 kilometers. Note that the decrease in risk of transmission per 100 kilometers is computed from Table 1 as (27% = 100% * [1 – exp(100 km * -0.0031)]). Beyond the effect of distance, the risk of transmission between individuals who reside within the same community was approximately 35-fold [13 - 98] higher at baseline and 8-fold [2 - 26] higher post-baseline compared to that between individuals residing in different communities. Note that the fold-change in the risk of transmission is computed from Table 1 as 35 = exp (3.56) at baseline and 8 = exp (2.05) post-baseline.

**Table 1:** Negative-binomial regression models describing the expected probability of viral linkage between a pair of individuals randomly sampled from their respective communities in the BCPP trial.

| Variable | Coefficient | Standard Error | 95% Conf. Interval | P value |
| --- | --- | --- | --- | --- |
| **Baseline model: Before the intervention had taken effect** |  |  |  |  |
| Intercept | −11.59 | 0.54 | −12.65 to −10.53 | < 0.001 |
| Transmission source: *control community* | 0.63 | 0.42 | −0.20 to 1.45 | 0.14 |
| Transmission type: *same community* | 3.56 | 0.52 | 2.54 to 4.59 | < 0.001 |
| Drive distance between communities in kilometers | −0.0025 | 0.0012 | −0.0047 to −0.0002 | 0.03 |
|  | **AIC** | 215.98 |  |  |
|  | **N** | 870 |  |  |
| **Post baseline model: After the intervention had taken effect** |  |  |  |  |
| Intercept | −11.31 | 0.54 | −12.37 to −10.24 | < 0.001 |
| Transmission source: *control community* | 0.90 | 0.51 | −0.11 to 1.90 | 0.08 |
| Transmission type: *same community* | 2.05 | 0.61 | 0.86 to 3.25 | 0.001 |
| Drive distance between communities in kilometers | −0.0031 | 0.0014 | −0.0059 to −0.0003 | 0.03 |
|  | **AIC** | 137.26 |  |  |
|  | **N** | 870 |  |  |

**Notes:**

**Figure 1.**
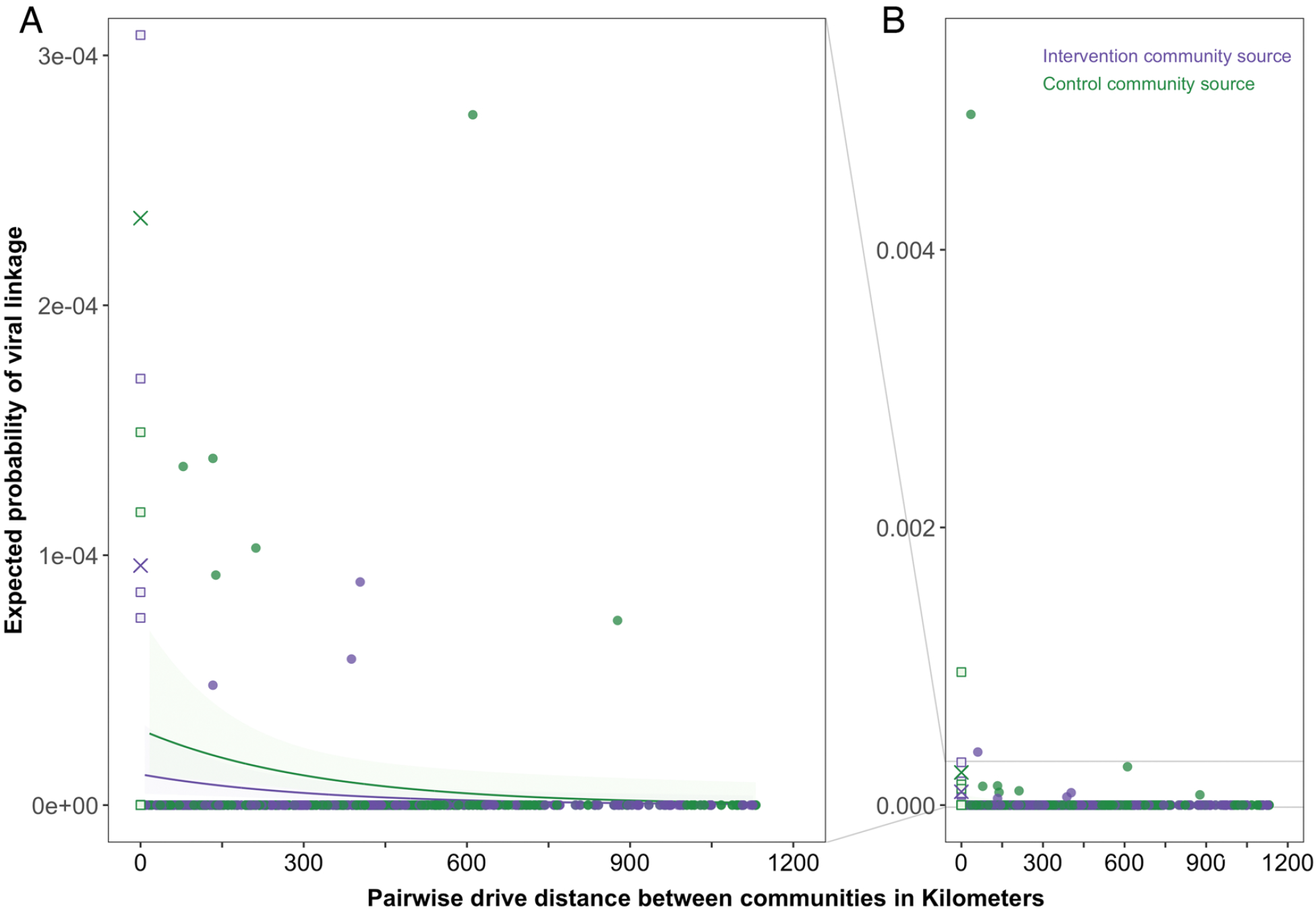
The risk of HIV-1 transmission between communities in the BCPP trial decreases as the drive distance separating them increases. The plot shows the expected probability of viral linkage, that is, risk of transmission between a pair of individuals randomly sampled from their respective communities in the BCPP trial. The expected probability of viral linkage was predicted with the post-baseline model in Table 1. To improve visibility **panel A** is a zoomed-in plot of the plot in **panel B**. Estimates for intervention community sources are shown in purple and those for control community sources are depicted in green. The solid curves and ribbons show the risk of transmission predicted by the post-baseline model between different communities in the BCPP trial and the associated uncertainty in the estimates. By comparison, the solid crosses depict the risk of transmission predicted by the post-baseline model within the same community. The squares and filled circles show the raw data for the 870 ordered community pairs of the 30 BCPP trial communities that were used to predict the expected probability of viral linkage within the same community (squares) or between different communities (filled circles). There were 15 intervention communities and 15 control communities in the BCPP trial. Because the “Digawana” intervention community had no participants with successfully sequenced samples during the post baseline period, community pairs with “Digawana” as a destination (recipient) community were excluded from the model (870 = 30 × 30 - 30). Among the 870 ordered community pairs, 14 were same community pairs with an intervention community as the origin (source) of transmission; 15 were same community pairs with a control community as the source of transmission, 421 were different community pairs with an intervention community as the source of transmission and 420 were different community pairs with a control community as the source of transmission. For each of the 870 ordered community pairs, the probability of viral linkage was computed from the raw data as the proportion of directed opposite-sex transmission pairs identified out of the total possible distinct opposite-sex transmission pairs among sampled participants.

### Estimating the relative contribution of different sources of infection residing inside versus outside the trial area

Next, using this model to estimate transmissions from communities that were not in the trial, we estimate proportions of transmissions into intervention communities and control communities of the BCPP trial that occurred from individuals in the same community; different communities in the same trial arm; different communities in the opposite trial arm; and non-trial communities (see “Supplementary Appendix sections S1.1 and S1.2”). We define non-trial communities as communities outside of the 30 communities that participated in the BCPP trial. We estimated that individuals in non-trial communities accounted for most of the transmissions that occurred to recipients in trial communities, with point estimates ranging from 84% to 92% in intervention communities and 73% to 92% in control communities (Figure 2). On average, 90% [95% Confidence Interval (CI): 81 – 93] of transmissions to recipients in intervention communities and 86% [74 – 90] of transmissions to recipients in control communities were estimated to have sources who lived in non-trial communities (Figure 3). This finding is consistent with communities in the BCPP trial being densely surrounded by communities outside the trial area and aligns with the fact that the BCPP trial participants represented a relatively small (7.6%) proportion of the national population.

**Figure 2.**
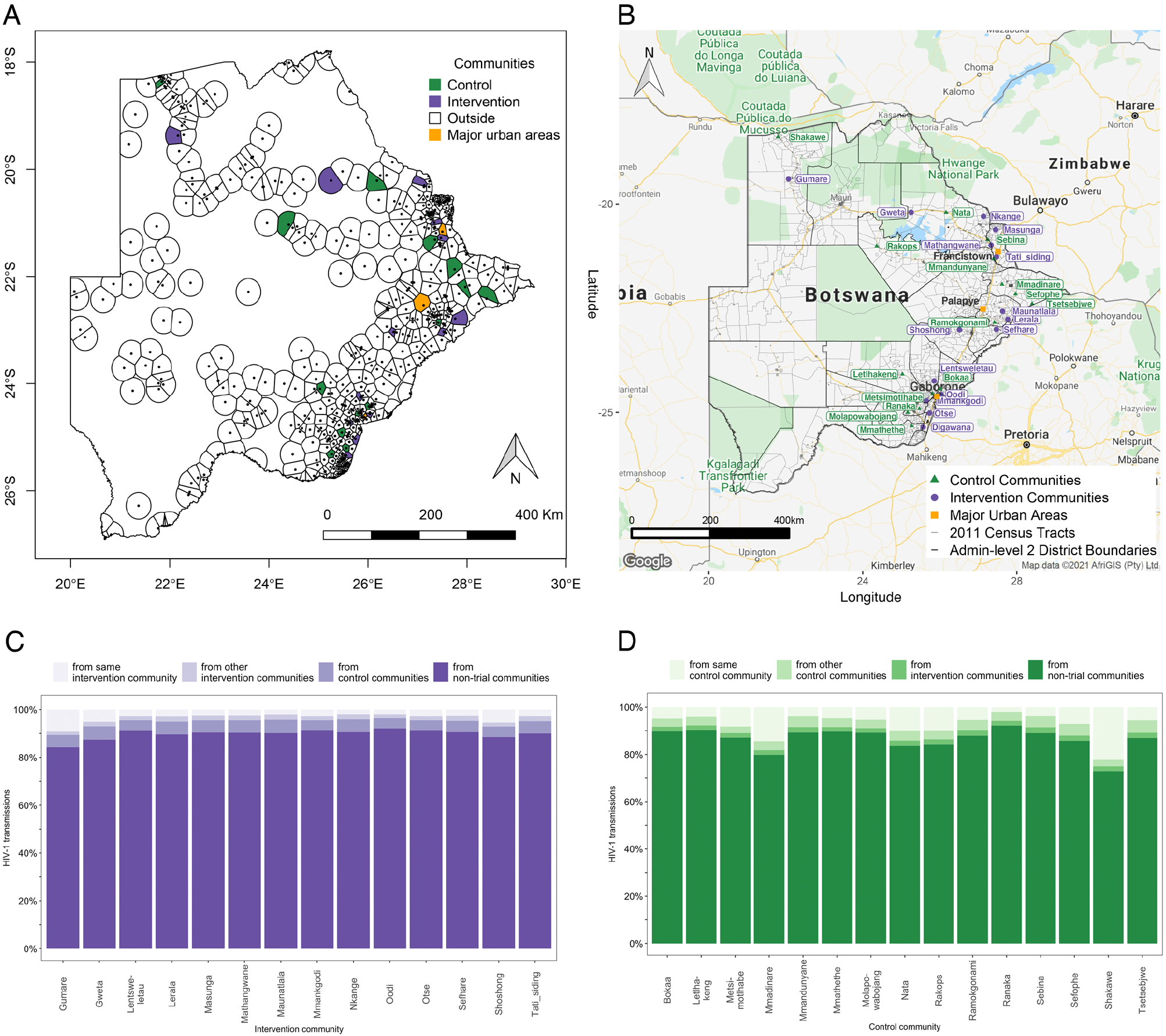
Estimated HIV-1 transmissions into communities in the BCPP trial from different sources of infection. **Panel A**. A voronoi tesselation map of communities (n = 488) in the 2011 Botswana population and housing census showing that communities in the BCPP trial are densely surrounded by communities outside the trial area i.e. non-trial communities. To complement Panel A, **Panel B** shows the names of the intervention communities and control communities in the BCPP trial within the context of administrative districts and census tracts. **Panel C** shows, in increasing shades of purple, the estimated proportions of HIV-1 transmissions to recipients in intervention communities from individuals in: the same community, other intervention communities, control communities and from non-trial communities. Note that “Digawana” intervention community is omitted from panel C because there were no successfully sequenced post-baseline samples in the community. **Panel D** shows, in increasing shades of green, the same for recipients in control communities. Most of the transmissions to recipients in BCPP trial communities originated from individuals in non-trial communities.

**Figure 3.**
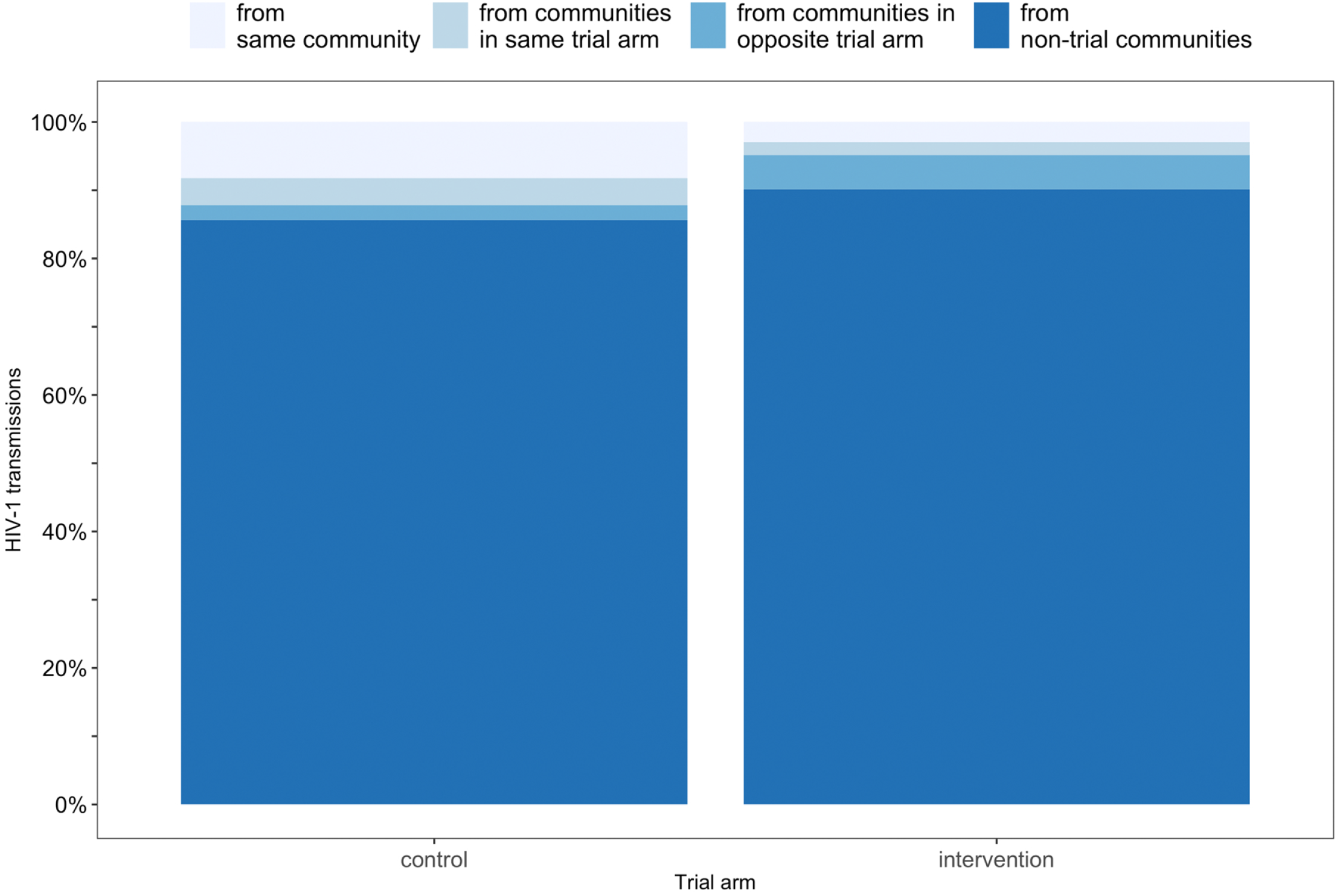
Mean estimates of HIV-1 transmissions that occurred to recipients in intervention communities and control communities in the BCPP trial from different sources of infection. The barplots show, in increasing shades of blue, the estimated proportions of HIV-1 transmissions to recipients in intervention communities and control communities from individuals in the same community (intervention: 2.9% [95% CI: 0.8− 10.4], control: 8.2% [7.6− 24.9]), communities in the same trial arm (intervention: 1.9% [0.6− 4.4], control: 4.0% [3.1− 4.3]), communities in the opposite trial arm (intervention: 5.0% [4.5− 5.2], control: 2.2% [0.7− 5.2]), and from non-trial communities (intervention: 90.1% [81.1− 93.1], control: 85.6% [73.6− 90.5]). The mean estimate of the proportion of HIV-1 transmissions to recipients in intervention communities from intervention sources, that is, from individuals in the same intervention community and from individuals in other intervention communities was 4.9% [95% CI: 1.7− 14.4].

### Proximity to urban centers

Communities in the BCPP trial are distributed around three major urban areas that each have relatively high numbers of people with HIV; these are Gaborone city in the South-East, Palapye in the Central-East and Francistown city in the North/North-East (Figure 2 and Supplementary Figure 2). Figure 2 shows that sexual partners in the same community had a greater impact on transmission in rural communities that are geographically isolated compared to in communities that closely neighbor major urban centers. For example, Gumare intervention community and Shakawe control community in the Northern region of Botswana received an estimated 9% [2 – 30] and 22% [8 - 55] of transmissions respectively from individuals in the same community; this estimated percentage was lower for communities on the periphery of densely populated urban areas such as Oodi intervention community 2% [0.4 – 7] and Bokaa control community 5% [2 – 15] in the South-East region. Furthermore, we found that the proportions of transmissions to recipients in intervention communities from individuals in the same trial arm were similar across the three major urban areas (Central-East: 5% [2 – 14], North/North-East: 6% [2 – 16], South-East: 4% [1 – 14]) (*χ*^2^ = 0.8, *df* = 2, *P*= 0.7).

### Impact of communities in the opposite trial arm

Individuals in control communities contributed a higher proportion of transmissions to intervention communities than the reverse. For example, the proportions of transmissions to recipients in intervention communities from individuals in control communities ranged with point estimates from 4.2% to 5.6%, compared to those to recipients in control communities from individuals in intervention communities that ranged from 1.9% to 2.4% (Figure 2). On average, 5.0% [4.5 – 5.2] of transmissions to recipients in intervention communities occurred from individuals in control communities compared to 2.2% [0.7 – 5.2] of transmissions to recipients in control communities that occurred from individuals in intervention communities, consistent with a benefit of treatment-as-prevention (Figure 3). Furthermore, Figure 3 shows that on average 2.9% [0.8 – 10.4] of transmissions to recipients in intervention communities occurred from individuals in the same community and 1.9% [0.6 – 4.4] of transmissions occurred from individuals in other intervention communities. In comparison, 8.2% [7.6 – 24.9] of transmissions to recipients in control communities occurred from individuals in the same community and 4.0% [3.1 – 4.3] of transmissions occurred from individuals in other control communities.

### Impact of a nationwide intervention

To evaluate what the impact of a national rollout of our HIV combination prevention intervention would have been on recipients in trial communities, we estimated what the number of transmissions would have been to recipients in trial communities if all (compared to none) of the non-trial communities nationwide had also received the intervention (see Supplementary Appendix section S1.2 “Counterfactual estimates”). We found that transmissions to recipients in trial communities could have been reduced by 59% [3 – 87] if the intervention had been applied nationally (Figure 4). Note that the proportion of transmissions in trial communities that could have been averted with a national rollout is computed from modeled estimates of the total number of transmissions to recipients in trial communities from sources inside trial communities as well as from sources in communities outside the trial area (averted transmissions = 100% * [(# transmissions into trial communities from all sources without intervention - # transmissions into trial communities from all sources with intervention) / # transmissions into trial communities from all sources without intervention]). Furthermore, this finding adds evidence that the impact of the BCPP trial intervention could be substantially larger than that observed in the trial if applied nationally.

**Figure 4.**
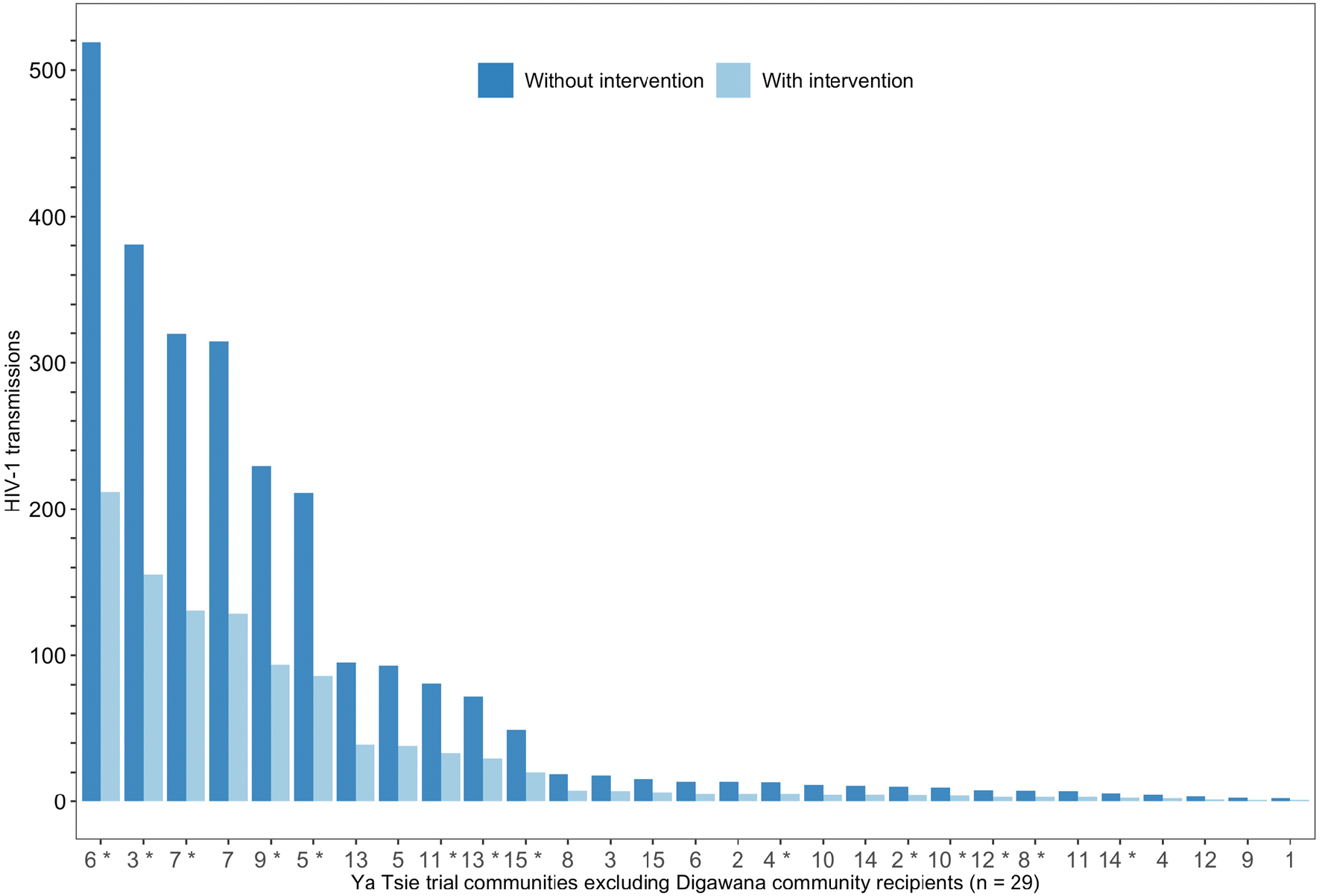
Counterfactual estimates of HIV-1 transmissions into BCPP trial communities showing the impact of a nationwide intervention. The grouped barplot shows the estimated number of transmissions to recipients in trial communities in the presence and absence of a nationwide intervention. Among the BCPP trial communities shown intervention communities are distinguished from control communities with an asterisk. The BCPP trial matched communities into 15 pairs based on geographical proximity to major urban areas, population-size and age structure, and access to health services. On average, a nationwide intervention could have reduced transmissions to recipients in trial communities by 59% [95% CI: 3− 87]. “Digawana” intervention community was excluded because there were no successfully sequenced post-baseline samples in the community that met inclusion criteria for phylogenetic analysis.

## Discussion

Global targets set by the Joint United Nations Programme on HIV/AIDS (UNAIDS) to have fewer than 500,000 new infections by the year 2020 on a path to reach epidemic control by the year 2030 were missed. Relevant to this, several large community-randomized universal HIV test-and-treat trials that were at the center of HIV prevention efforts in East and Southern Africa showed mixed results [1-5]. To aid interpretation of the complex trial results and inform public health policy decisions about effective HIV prevention strategies, we developed a statistical modeling approach that uses directed sexual contacts inferred from deep-sequenced HIV virus to quantify the relative contribution of different sources of infection, that might be inside trial communities or outside the trial area. Briefly, to demonstrate the relative extent to which transmissions in intervention communities and control communities of the BCPP trial in Botswana occurred from individuals in the same community; different communities in the same trial arm; different communities in the opposite trial arm; and communities outside the trial area we first inferred directed opposite-sex transmission events between trial communities using deep-sequence phylogenetics. Then we used the directed opposite-sex transmission events that were inferred from deep-sequence phylogenetics together with the pairwise drive distances between trial communities and the intervention status of source communities to statistically model the risk of transmission between trial communities. After that we provided pairwise drive distances between communities that participated in the 2011 Botswana population and housing census to the model as input to estimate the risk of transmission (expected probability of viral genetic-linkage had the cases been sequenced) between communities nationally. Then, to estimate the number of transmissions into trial communities from all communities nationally we combined estimates of the risk of transmission to recipients in trial communities from communities nationally with population-size estimates from the 2011 Botswana population and housing census [14] and district-level HIV prevalence estimates from the 2013 Botswana AIDS Impact Survey (BAIS 2013) [15].

Power analyses and model predictions for the primary endpoints of the BCPP trial in Botswana and the PopART trial in South Africa and Zambia assumed that 20% [95% CI: 15 – 25] and 5% of sexual partnerships would involve a partner outside one’s own community, respectively [2, 8, 16]. Strikingly, we found that individuals in non-intervention communities accounted for most of the transmissions that occurred to recipients in intervention communities; with an estimated 90% [81 – 93] of transmissions attributable to individuals from non-trial communities and 5.0% [4.5 – 5.2] of transmissions attributable to individuals from control communities. For context, a phylogenetic study that used consensus sequences of the HIV-1 *POL* (polymerase) gene to estimate the relative contribution of local transmission versus external introductions to HIV-1 incidence in the Africa Health Research Institute (AHRI) study population, a rural and peri-urban population located immediately adjacent to the TasP trial study area in KwaZulu-Natal, South Africa, estimated that 35% [20 – 60] of new infections in the study population were external introductions that occurred from sexual partners outside the study area [5, 17]. Most of the external introductions in the AHRI phylogenetics study were estimated to be from sources within the national borders of South Africa with few cross-border external introductions from Botswana, Malawi, Mozambique, Zambia and Zimbabwe (see Figure 2B in [17]). BCPP trial communities closely neighbor three major urban areas in Botswana (Gaborone city in the South-East, Palapye in the Central-East and Francistown city in the North/North-East), and people tend to be fairly mobile in Botswana. By comparison, the AHRI study area is relatively distantly located from the major urban area in the KwaZulu-Natal province of South Africa (200 kilometers north of Durban city), therefore, it is unsurprising that there would be more external introductions to BCPP trial communities compared to the AHRI study population. In line with the finding from the AHRI phylogenetics study, a clustering analysis conducted by the PANGEA-HIV consortium on HIV-1 viral consensus sequences from the AHRI study population in South Africa, BCPP trial in Botswana, MRC study population in Uganda, PopART study population in Zambia and Rakai study population in Uganda found few clusters including cohorts from different countries. The limited cross-border external introductions into Botswana suggest that extending the BCPP trial intervention to all communities nationally to target more sources could effectively reduce the occurrence of new infections.

Accordingly, we estimated in a counterfactual modeling scenario to demonstrate the impact of a national application of the intervention that applying the BCPP trial intervention in all communities nationwide (versus none) could have reduced transmissions to recipients in trial communities by 59% [3 – 87]) at the time (compared with the 30% reduction that was observed in the BCPP HIV incidence cohort). This was done under an assumption that the intervention effect that was observed in intervention communities would be similar when extended to a larger geographical area and would reduce the risk of transmission between any two individuals residing in Botswana, rather than (as in the trial) any two individuals residing in intervention communities. In practice, the intervention effect could vary owing to differences in transmission patterns of different population sub-groups and geographical locations. These findings suggest that substantial reductions in transmission could be achieved if the intervention is applied nationwide and that estimating the relative contribution of various sources of transmission (attributable fraction of cases) could help to guide targeted applications of the intervention where resources are limited. The timing of the implementation of a universal test-and-treat intervention could be crucial. Fast roll-out could limit the spread of infection and shorten the time to reach epidemic control. Furthermore, the estimated reductions in transmissions with a nationwide intervention suggest that the universal HIV test-and-treat intervention could be used as a foundation for incidence reduction upon which other interventions could be layered to close the gap to reach epidemic control.

A key strength of our statistical approach is that we demonstrate how deep-sequence pathogen genomics can be used at scale to assess interventions in cluster-randomized trials of infectious disease prevention. Our analysis is based on the central assumptions that transmission patterns in communities randomized to the control arm of the trial are representative of those found in non-trial communities, and that, the population-size and HIV prevalence of communities are known; and that the HIV prevalence in administrative districts is representative of that in communities (see Supplementary Appendix section S1.1 “Population-based molecular source attribution model”). There are some limitations to our analysis: First, our statistical approach is informed by pairwise drive distances separating pairs of communities and could be improved with mobile phone data to gain insight on daily commutes and seasonal migration for work (for example: farming and mining) and holidays. Second, HIV viral sequences of cases were collected only in trial communities. However, Figure 1 and Table 1 show a relationship between the drive distance separating pairs of communities in the BCPP trial and the estimated risk of transmission between them. This relationship allows us to use the drive distances separating trial communities and non-trial communities to estimate the expected probability of viral linkage to source cases in non-trial communities had cases in non-trial communities also been sequenced. Third, even though we did not explicitly model the impact of community size on risk of HIV-1 transmission to- and from-communities we found that there was generally a positive correlation between the number of transmissions to recipients in trial communities predicted by the post-baseline model in Table 1 and the opportunity for transmission to recipients in trial communities. The opportunity for transmission to recipients in a community is defined as the maximum distinct possible opposite-sex transmission pairs that could involve recipients in that community and is based on the number of people with HIV in the source and recipient communities (Supplementary Figure 4). To broaden insights, our statistical modeling approach could be applied to estimate the relative contribution of various sources of infection by age and sex in the other community-randomized universal HIV test-and-treat trials that have assembled deep-sequence genomic data, for example the PopART trial in South Africa and Zambia.

Our findings have implications for public health policy and for the design of effective HIV prevention strategies. By deconstructing the relative contribution of different sources of infection in intervention communities versus control communities this work aids interpretation of the complex universal HIV test-and-treat trials in which the intervention is administered on one group (people with HIV) and the outcome (reduction in number of new cases) is measured on another group (people in the same community without HIV). For example, our findings elucidate the potential impact of a nationwide intervention and provide insight on the extent to which the BCPP intervention was diluted by spillover infections from control communities and from communities outside the trial area. Furthermore, our findings inform on-going public health policy discussions on whether the HIV testing component in national HIV prevention programs should be centered on facility-based testing at clinics and index-based testing of family and sexual contacts of people with HIV or anchored on intensive universal household-based HIV testing as was done in the trials. For example, this study shows how the combination of universal household-based HIV testing and routine HIV testing in health facilities - as was done in the combination prevention intervention in the BCPP trial - allows us to infer transmission patterns within and between communities to guide HIV prevention strategies.

In sum, this work shows that individuals residing in communities outside the BCPP trial area accounted for most of the transmissions to recipients in intervention communities, limiting the impact of the BCPP trial intervention. Furthermore, substantial gains in reducing transmission could be made with a nationwide application of the intervention. With the introduction of interventions at the community-level (universal test-and-treat) and individual-level (pre-exposure prophylaxis and self-testing) our analysis suggests that genomic surveillance could provide a crucial platform to assess interventions allowing us to track how pathogens spread and evolve overtime in response to different interventions. For example, samples collected for routine viral load testing could also be sequenced to track the directional spread of infection within and between communities and between age-sex population sub-groups. This could help to identify population sub-groups and communities in which HIV prevention interventions need to be further strengthened to reduce transmission. Pairing genomic information with information from studies that explicitly quantify the impact of social behavioral change on interventions could aid the interpretation of HIV prevention studies and evidence-based policy design. Based on our findings, we recommend that studies of infectious disease prevention consider the impact of sources of transmission beyond the reach of the intervention when evaluating interventions to inform public health programs.

## Data Availability

All relevant data are within the paper, figures and tables. A code repository has been made available at the following URL: https://github.com/magosil86/spillover-infections

## Supplementary Appendix S1 Statistical approach to estimate the relative contribution of various sources of infection in cluster-randomized trials of infectious disease prevention

### S1.1 Population-based molecular source attribution model

In cluster-randomized trials of HIV prevention, where the randomization unit is communities, new infections can arise from individuals: in the same community, in different communities in the same trial arm, in different communities in the opposite trial arm and in non-trial communities. The aim of this analysis is to estimate the proportions of transmissions in trial communities that are attributable to the above-mentioned sources of infection. Suppose we would like to estimate the number of transmissions in the population, *z*_*ij*_ that occurred to individuals in community *i* from individuals of the opposite-sex in community *j*. We refer to community *i* as a recipient community and community *j* as a source community. Notice that, *i = j* when estimating the number of transmissions that occurred from individuals in the same community. We estimate the number of transmissions into a recipient community *i* from a source community *j* at period *t* using two quantities, *NH*_*ij*_(*t*) which is treated as known and represents the maximum number of distinct possible (opposite-sex) transmission pairs in the population between the two communities at period *t* and, *π*_*ij*_(*t*) the risk of transmission between the two communities at period *t*, that is, the expected probability of viral-linkage between deep-sequenced HIV viruses of individuals randomly sampled from the respective communities. More precisely, we consider the model

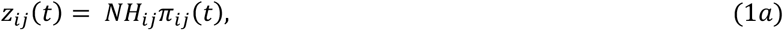

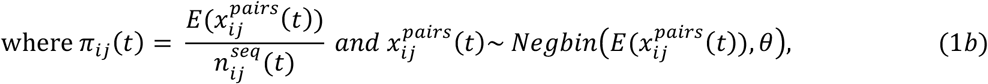

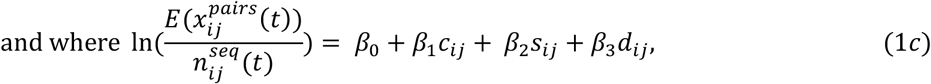

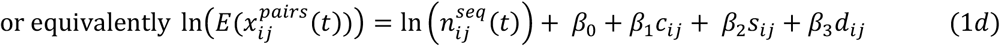

with data and quantities

*N*_*i*_ population-size of community *i*

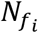 female population-size of community *i*

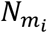 male population-size of community *i*

*N*_*j*_ population-size of community *j*

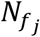 female population-size of community *j*

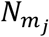 male population-size of community *j*

*H*_*i*_ HIV prevalence of community *i*

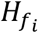 female HIV prevalence of community *i*

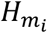 male HIV prevalence of community *i*

*H*_*j*_ HIV prevalence of community *j*

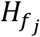 female HIV prevalence of community *j*

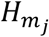 male HIV prevalence of community *j*

*NH*_*i*_ = *N*_*i*_ * *H*_*i*_ number of people with HIV in community *i*

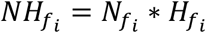 number of females with HIV in community *i*

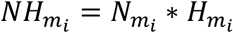 number of males with HIV in community *i*

*NH*_*j*_ = *N*_*j*_ * *H*_*j*_ number of people with HIV in community *j*

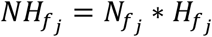 number of females with HIV in community *j*

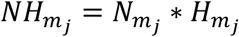 number of males with HIV in community *j*

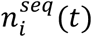 number of individuals randomly sampled from community *i* at period *t* whose viral whole genomes were successfully deep-sequenced

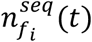 number of females randomly sampled from community *i* at period *t* whose viral whole genomes were successfully deep-sequenced

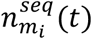 number of males randomly sampled from community *i* at period *t* whose viral whole genomes were successfully deep-sequenced

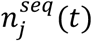 number of individuals randomly sampled from community *j* at period *t* whose viral whole genomes were successfully deep-sequenced

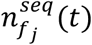 number of females randomly sampled from community *j* at period *t* whose viral whole genomes were successfully deep-sequenced

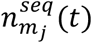 number of males randomly sampled from community *j* at period *t* whose viral whole genomes were successfully deep-sequenced

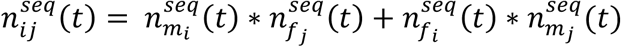 maximum number of distinct possible (opposite-sex) transmission pairs at period *t* between individuals randomly sampled from communities *i* and *j* and whose viral whole genomes were successfully deep-sequenced

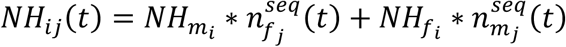 maximum number of distinct possible (opposite-sex) transmission pairs in the population at period *t* between communities *i* and *j*

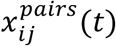 number of transmission pairs identified from the deep-sequenced HIV virus of individuals randomly sampled from communities *i* and *j* at period *t*

*c*_*ij*_ source community is a nonintervention community, that is, a control community or non-trial community (yes = 1, no = 0)

*s*_*ij*_ source community and recipient community are the same, that is, same community transmission (yes = 1, no = 0)

*d*_*ij*_ distance in kilometers separating the source community and recipient community and estimated parameters

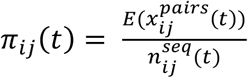 risk of HIV transmission between communities *i* and *j* at period *t*

*θ* overdispersion parameter

*β*_0_, *β*_1_, *β*_2_, *β*_3_ fixed effects regression parameters

For clarity, the maximum number of distinct possible (opposite-sex) transmission pairs between individuals randomly sampled from communities *i* and *j* during the baseline period of the trial is, 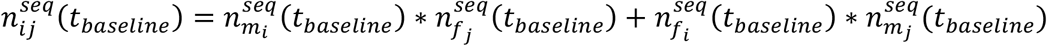. By comparison, the maximum number of distinct possible (opposite-sex) transmission pairs between individuals randomly sampled from communities *i* and *j* during the post-baseline period of the trial is, 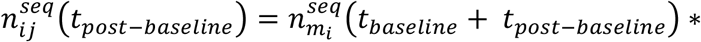 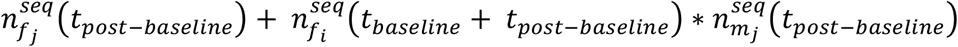. The central assumptions of this model are that the transmission pairs identified between samples from communities *i* and *j* are independent, and that transmission patterns in communities randomized to the control arm of the trial are representative of those found in non-trial communities. The simplifying assumption of independence is appropriate when identified transmission events mostly comprise small two-person clusters as found in the BCPP data and could be less-suited for large clusters typical of super spreader events. Furthermore, we assume that population-size and HIV prevalence of communities *i* and *j* are known, and that the HIV prevalence in administrative districts is representative of that in communities. We acknowledge that community-level HIV prevalence estimates can be obtained directly with the methods of [19] and reserve such computation for future study.

### S1.2 Application to BCPP study data

We applied the population-based molecular source attribution model in section S1.1 to estimate the relative contribution of sources of infection (inside versus outside the trial area) to transmissions that occurred to recipients in BCPP trial communities. Because viral genotyping was only done in trial communities, we used census data to combine information from trial communities with that from communities outside the trial area. First, we estimated the maximum number of distinct possible (opposite-sex) transmission pairs in the population between ordered pairs (488 × 29) of the 488 communities that participated in the 2011 Botswana population and housing census (see methods “*Pairwise drive distance data*” and “*Population-size and HIV prevalence estimates*”). Because there were no individuals sampled from “Digawana” intervention community during the post-baseline period whose HIV-1 virus was successfully deep-sequenced and met inclusion criteria for phylogenetic analysis, we excluded ordered community pairs that had “Digawana” as a recipient (destination) community. Afterwards, as described in equations 1b to 1d, we used directed opposite-sex transmission pairs identified between ordered pairs of the 30 communities in the BCPP trial (Supplementary Figure 1) to estimate the risk of transmission, that is, expected probability of viral linkage between those ordered community pairs during the baseline and post-baseline periods of the trial (see Table 1, Figure 1 and methods “*Deep-sequence phylogenetics data*” and “*Pairwise drive distance data*”). Estimates were obtained using parametric maximum likelihood estimation with the nbreg module in Stata 13.1 and the glm.nb function in the MASS package v7.3-54 in R v4.1.2 [13]. We excluded ordered community pairs that had “Digawana” as a recipient (destination) community from the input datasets used to model the risk of transmission for the same reasons as described above, in particular, there were no individuals sampled from “Digawana” intervention community during the post-baseline period whose HIV-1 virus was successfully deep-sequenced and met inclusion criteria for phylogenetic analysis.

Consequently, the input datasets used to fit the baseline and post-baseline models to estimate the risk of transmission between communities in the BCPP trial comprised 870 distinct observations (870 = 30 × 30 - 30) that each contained six pieces of information: 1) ordered community pair, that is, source community and recipient community, 2) non-intervention community status, 3) same community transmission status, 4) drive distance in kilometers separating the source community and recipient community, 5) number of transmission pairs identified (observed) between individuals randomly sampled from the source community and recipient community during the relevant time period, and 6) the maximum number of distinct possible (opposite-sex) transmission pairs between individuals randomly sampled from the source community and recipient community during the relevant time period. Next, we used the post-baseline model of the risk of transmission between ordered pairs of communities in the BCPP trial to predict the risk of transmission to recipients in intervention communities and control communities from the 488 census communities. The risk prediction dataset comprised (14,152 = 488 × 29) distinct observations that each contained five pieces of information. These were: the same first four pieces of information as those in the input dataset for the post-baseline model of the risk of transmission, and for the fifth piece of information, we set the natural-log of the maximum number of distinct possible (opposite-sex) transmission pairs between individuals randomly sampled from the source community and recipient community during the relevant time period to zero, that is, 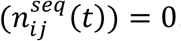. We set 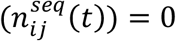to predict the risk of transmission (expected probability of viral linkage) instead of the expected transmission counts. In each ordered community pair, we used the origin community in an origin-destination pairing as a surrogate for the source community and the destination community as a surrogate the recipient community. Predictions were made with the predict function in the stats package in R v4.1.2. Then, as described in equation 1a, we estimated the number of transmissions between each ordered pair of communities as the product of the maximum number of distinct possible (opposite-sex) transmission pairs in the population between the ordered community pairs and the estimated risk of transmission between them.

After estimating the number of transmissions that occurred at period *t* between all ordered community pairs in the population that have community *i* as a recipient community, we denote the total estimated number of transmissions to recipients in community *i* at period *t* as the vector

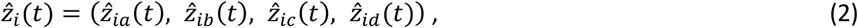

where 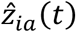 is the estimated number of transmissions from individuals within the same community, that is, community *i*, 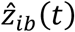 is the estimated number of transmissions from individuals in other communities that are in the same trial arm as community *i*, 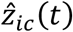 is the estimated number of transmissions from individuals in communities that are in the opposite trial arm to community *i* and 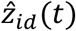 is the estimated number of transmissions from individuals in non-trial communities. Then we estimate the proportions of transmissions to recipients in community *i* from individuals (sources of infection) in different types of communities as

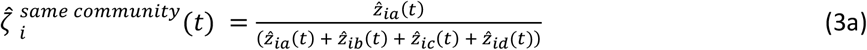

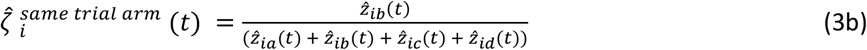

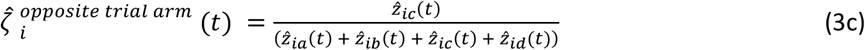

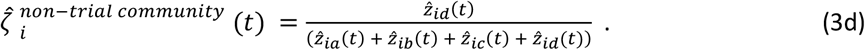

Suppose community *i* was randomized to receive the intervention then the mean estimate of the proportion of transmissions to recipients in intervention communities from individuals in the same community is

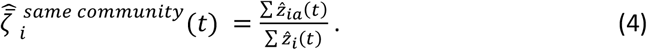

The same principle follows for the three other sources of infection: same trial arm, opposite trial arm and non-trial community; wherein the numerator for the mean estimate is the sum of the numerators of the individual community estimates and similarly, the denominator for the mean estimate is the sum of the denominators of the individual community estimates (Figures 2 and 3).

#### Counterfactual estimates

To model the impact of a nationwide intervention (Figure 4) we estimated what the number of transmissions to recipients in trial communities would have been if all 488 census communities had received the intervention (*c*_*ij*_ = 0) compared to if none of the communities had received the intervention (*c*_*ij*_ = 1). This was done under an assumption that the observed effect of the BCPP trial intervention in intervention communities would be similar when extended to a larger geographical area. However, there could be variation in the effect of the intervention that is observed due to heterogeneous transmission patterns across different population sub-groups and geographical locations. Because there were no individuals sampled from “Digawana” intervention community during the post-baseline period whose HIV-1 virus was successfully deep-sequenced and met inclusion criteria for phylogenetic analysis we excluded ordered community pairs that had “Digawana” as a recipient (destination) community. We estimate that a nationwide application of the intervention could have reduced transmissions to recipients in trial community *i* by,

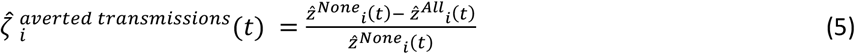

and that on average transmissions across all trial communities could have been reduced by,

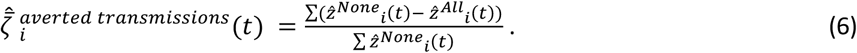

#### Alternative models

We considered alternative versions of the post-baseline model for the risk of transmission between ordered pairs of communities in the BCPP trial that used transforms of the pairwise drive distance separating communities (Supplementary Table 1). The linear model shown in supplementary Table 1 and described in equations 1b through 1d was selected as the best model based on Aikaike information criterion (AIC) and parsimony. We noted that sampling of trial participants through clinics in the BCPP trial was only done in intervention communities but not in control communities resulting in an asymmetry between the intervention and control arms of the trial. Therefore, we modeled the risk of transmission between ordered pairs of communities in the BCPP trial excluding transmission pairs where both individuals were sampled at a clinic in an intervention community during the same trial period, that is, both individuals sampled at baseline or post-baseline (Supplementary Table 2). We found similar patterns to those observed in the baseline and post-baseline models in Table 1. The negative-binomial regression models used to estimate the risk of transmission between ordered pairs of communities in the BCPP trial were fit with parametric maximum likelihood estimation with the nbreg module in Stata 13.1 and the glm.nb function in the MASS package v7.3-54 in R v4.1.2 [13].

#### Model diagnostics

We performed several diagnostics to assess the fit of the post-baseline model in Table 1 to the directed transmission pairs identified between ordered pairs of communities in the BCPP trial. First, we assessed if the model converged to the maximum likelihood of the data using a likelihood grid search wherein the mean number of transmission pair counts predicted by the post-baseline model in Table 1 was adjusted upwards and downwards by 10% and 20%. We found that adjusting the predicted number of counts upwards or downwards did not improve the log-likelihood suggesting that the model had converged on the maximum likelihood of the data. Second, we compared the number of transmission pairs identified between ordered pairs of communities in the BCPP trial with those that would be expected under the post-baseline model in Table 1. There was little evidence to suggest that the observed counts differed substantially from those expected under the model (Fisher exact *P* = 0.792) (Supplementary Table 3). Third, we also used a simulation-based approach to compare the distribution of observed quantile residuals with that expected under the post-baseline model in Table 1 and found little appreciable difference between the observed and expected distributions (Supplementary Figure 3). The simulation-based approach was performed using the DHARMa package v0.4.6 in R v4.1.2.

#### Confidence intervals

We used an empirical bootstrap approach to compute 95% confidence intervals for the estimated proportions of transmissions attributable to different sources of infection wherein each bootstrap procedure was performed with 1,000 replicates. The lower and upper bounds of the confidence intervals represent the 2.5% and 97.5% quantiles, respectively.

## Acknowledgements and Funding

We are grateful to participants and collaborators from the Botswana Combination Prevention Project for their support during this work. We also thank the following colleagues for their helpful suggestions: Stephanie Marie Davis, Carol A. Ciesielski, Anindya De and Stacie Greby. This study was supported by the National Institute of General Medical Sciences (U54GM088558); the Fogarty International Center (FIC) of the U.S. National Institutes of Health (D43 TW009610); and the President’s Emergency Plan for AIDS Relief through the Centers for Disease Control and Prevention (CDC) (Cooperative agreements U01 GH000447 and U2G GH001911), NIH K24 AI131928 as well as the Morris-Singer Fund, the VK Fund for the Harvard Center for Communicable Disease Dynamics and the Bill and Melinda Gates Foundation.

## Disclaimer

The findings and conclusions in this report are those of the author(s) and do not necessarily represent the official position of the funding agencies.

---

**Supplementary Table 1:** A comparison of three negative-binomial regression models that describe the expected probability of viral linkage between a pair of individuals randomly sampled from their respective communities in the BCPP trial. The models are fit ***(with and without) a transformation of the drive distance between communities***.

| Variable | Coefficient | Standard Error | 95% Conf. Interval | P value |
| --- | --- | --- | --- | --- |
| **Linear model** |  |  |  |  |
| Intercept | -11.31 | 0.54 | -12.37 to -10.24 | < 0.001 |
| Transmission source: *control community* | 0.90 | 0.51 | -0.11 to 1.90 | 0.08 |
| Transmission type: *same community* | 2.05 | 0.61 | 0.86 to 3.25 | 0.001 |
| Drive distance between communities in kilometers | -0.0031 | 0.0014 | -0.0059 to -0.0003 | 0.03 |
|  | **AIC** | 137.26 |  |  |
|  | **N** | 870 |  |  |
| **Model with log transformed drive distance** |  |  |  |  |
| Intercept | -8.23 | 1.36 | -10.90 to -5.57 | < 0.001 |
| Transmission source: *control community* | 0.96 | 0.53 | -0.08 to 2.01 | 0.07 |
| Transmission type: *same community* | -1.04 | 1.41 | -3.80 to 1.73 | 0.46 |
| Log<sub>e</sub>(drive distance between communities in kilometers) | -0.77 | 0.26 | -1.28 to -0.25 | 0.004 |
|  | **AIC** | 137.98 |  |  |
|  | **N** | 870 |  |  |
| **Model with squared drive distance** |  |  |  |  |
| Intercept | -11.11 | 0.63 | -12.35 to -9.88 | < 0.001 |
| Transmission source: *control community* | 0.89 | 0.51 | -0.11 to 1.90 | 0.08 |
| Transmission type: *same community* | 1.86 | 0.70 | 0.48 to 3.24 | 0.008 |
| Drive distance between communities in kilometers | -0.0046 | 0.0033 | -0.0110 to 0.0017 | 0.15 |
| Squared (drive distance between communities in kilometers) | $1.70E - 06$ | $2.75E - 06$ | $-3.69E - 06$ to $7.08E - 06$ | 0.54 |
|  | **AIC** | 139.03 |  |  |
|  | **N** | 870 |  |  |

**Notes:**

**Supplementary Table 2:**
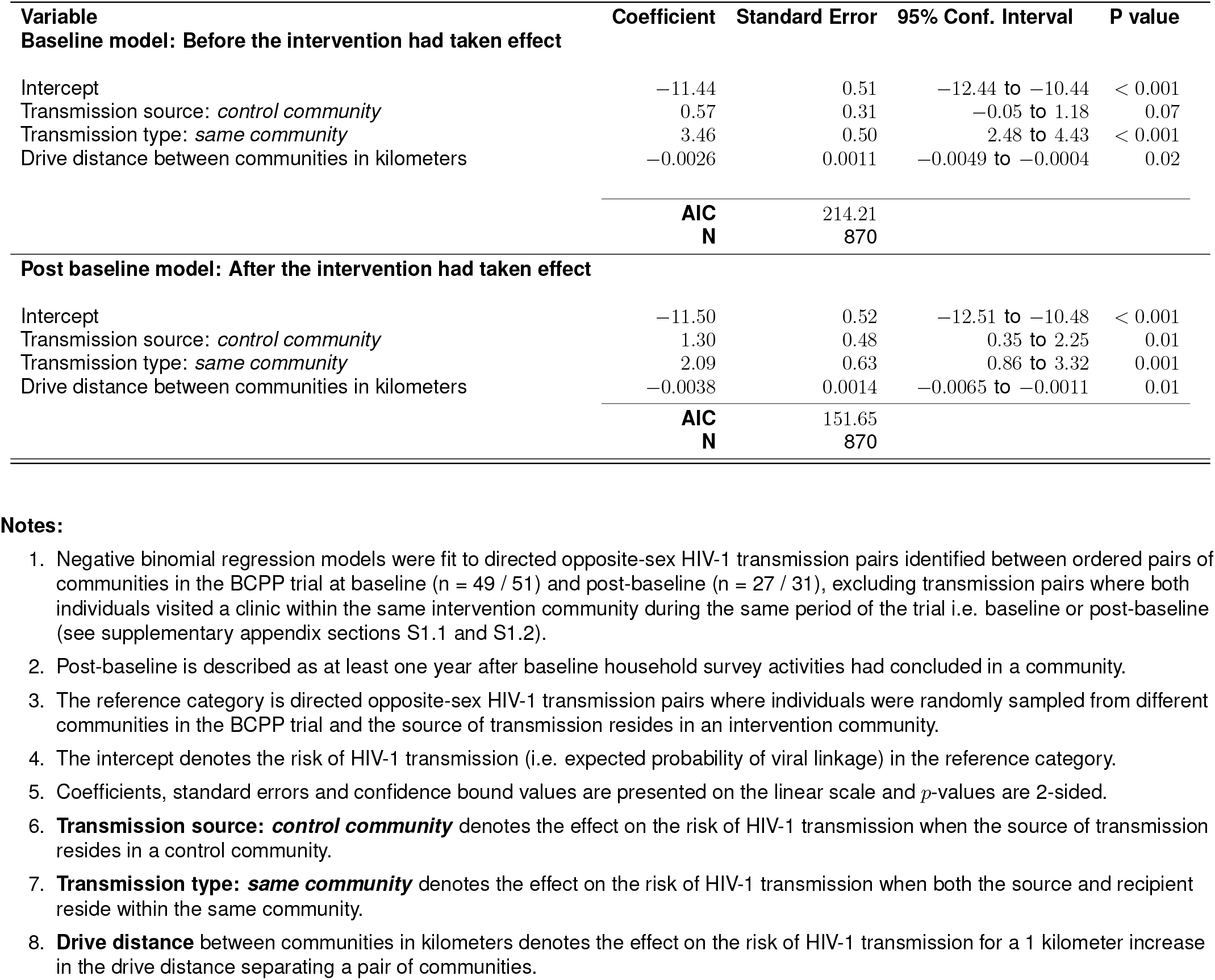
Negative-binomial regression models describing the expected probability of viral linkage between a pair of individuals randomly sampled from their respective communities in the BCPP trial. Compared with Table 1, the models ***exclude potential partner co-visit events to clinics in intervention communities during baseline or post-baseline***.

| Variable | Coefficient | Standard Error | 95% Conf. Interval | P value |
| --- | --- | --- | --- | --- |
| **Baseline model: Before the intervention had taken effect** |  |  |  |  |
| Intercept | −11.44 | 0.51 | −12.44 to −10.44 | < 0.001 |
| Transmission source: *control community* | 0.57 | 0.31 | −0.05 to 1.18 | 0.07 |
| Transmission type: *same community* | 3.46 | 0.50 | 2.48 to 4.43 | < 0.001 |
| Drive distance between communities in kilometers | −0.0026 | 0.0011 | −0.0049 to −0.0004 | 0.02 |
|  | **AIC** | 214.21 |  |  |
|  | **N** | 870 |  |  |
| **Post baseline model: After the intervention had taken effect** |  |  |  |  |
| Intercept | −11.50 | 0.52 | −12.51 to −10.48 | < 0.001 |
| Transmission source: *control community* | 1.30 | 0.48 | 0.35 to 2.25 | 0.01 |
| Transmission type: *same community* | 2.09 | 0.63 | 0.86 to 3.32 | 0.001 |
| Drive distance between communities in kilometers | −0.0038 | 0.0014 | −0.0065 to −0.0011 | 0.01 |
|  | **AIC** | 151.65 |  |  |
|  | **N** | 870 |  |  |

**Notes:**

**Supplementary Table 3:**
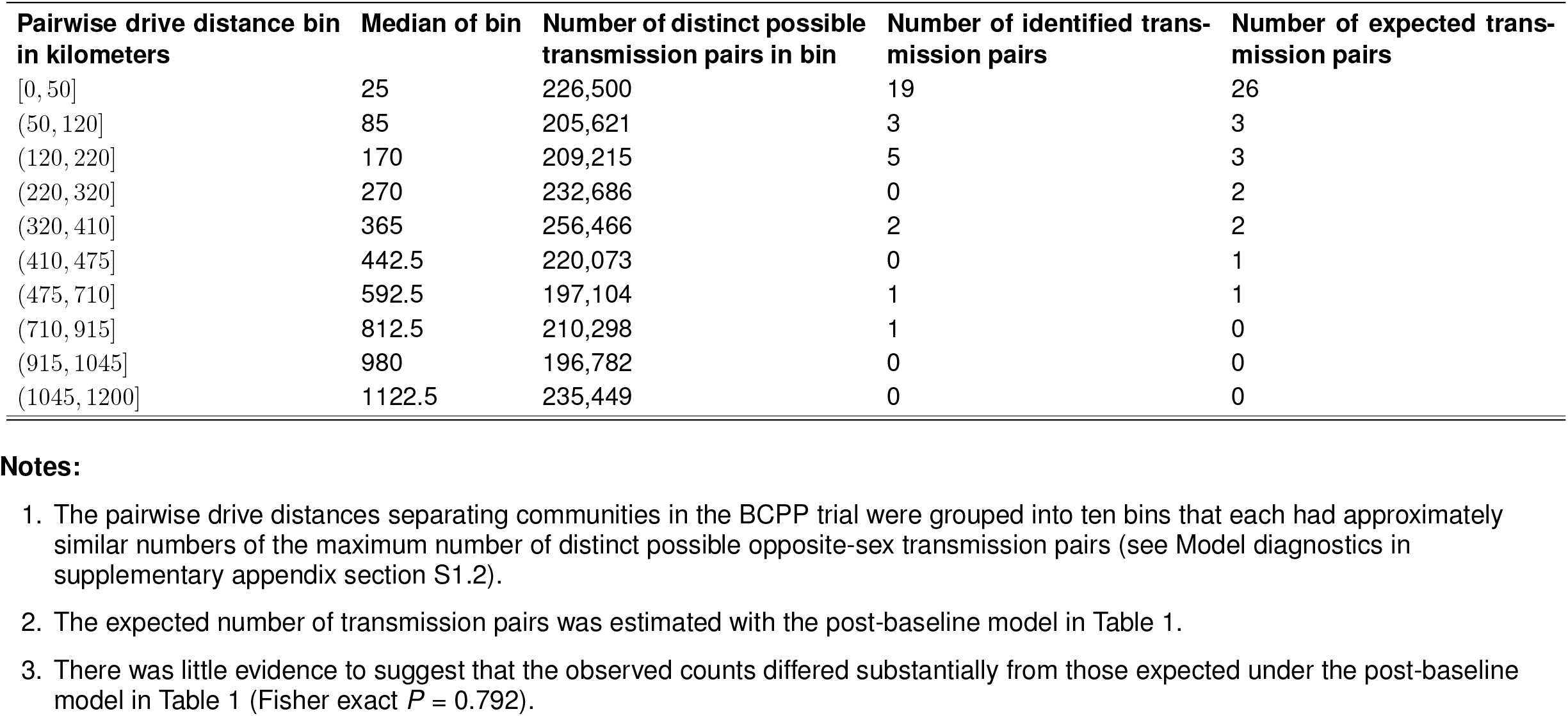
A comparison of the number of transmission pairs identified in the BCPP trial during the post-baseline period (n = 31) with those expected under the post-baseline model in Table 1. Table 1 describes the risk of HIV-1 transmission between communities.

**Supplementary Figure 1.**
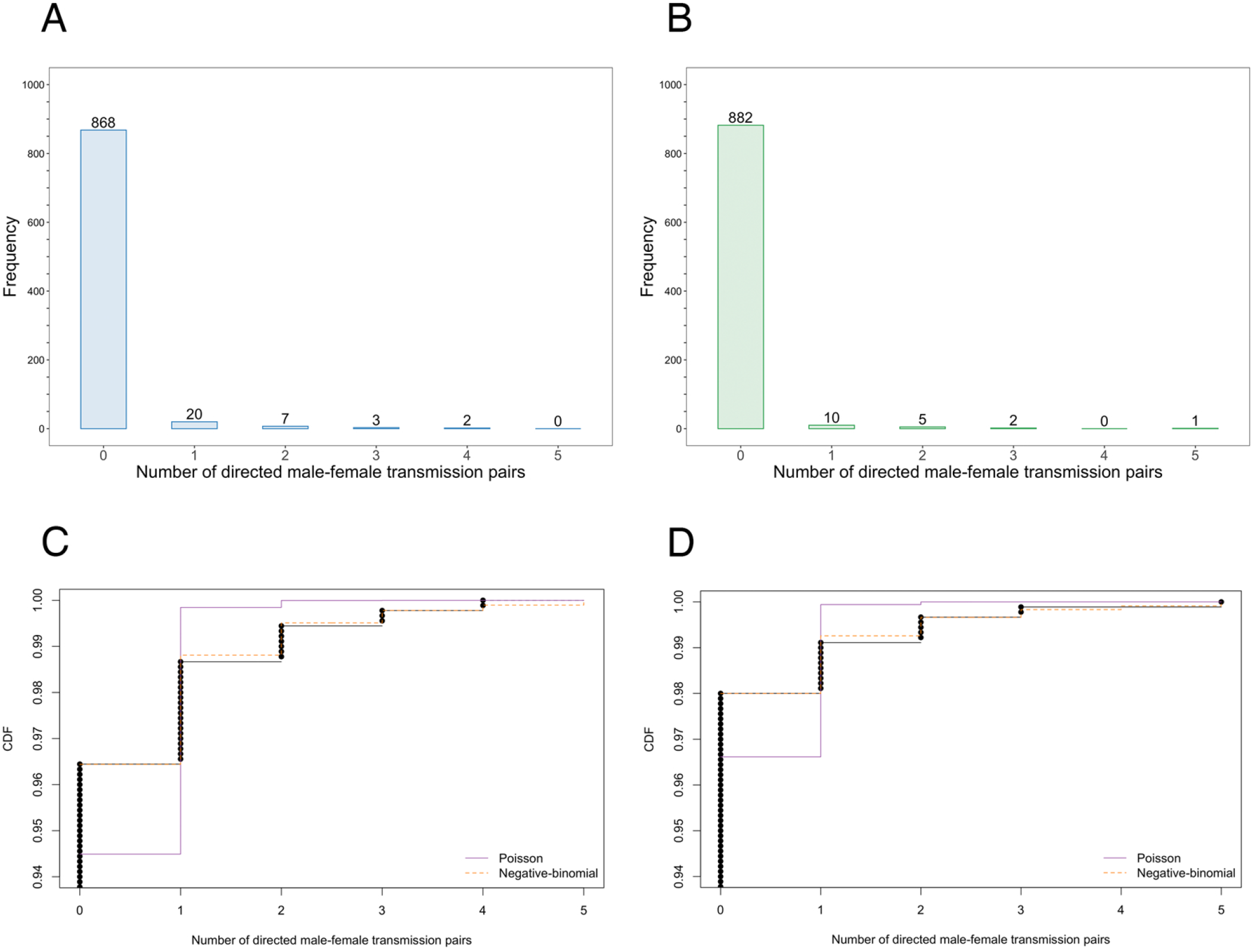
Distribution of directed opposite-sex transmission pairs identified between ordered pairs of the 30 communities in the BCPP trial. The barplots in panels **A** and **B** show distributions of transmission pairs identified during the baseline (blue bars) and post-baseline (green bars) periods of the BCPP trial, respectively. For example, in panel A there were 20 ordered community pairs that each had a single identified opposite-sex transmission pair and 7 ordered community pairs that each had 2 identified transmission pairs. Panels **C** and **D** show the corresponding empirical and theoretical cumulative distribution functions (cdf) of identified transmission pairs during the baseline **(panel C)** and post-baseline **(panel D)** periods of the BCPP trial. In both panels **C** and **D** the empirical distribution is shown in black, and the theoretical Poisson and negative-binomial distributions are illustrated by solid purple lines and orange broken lines, respectively. The positively skewed distributions of identified transmission pairs in the BCPP trial are better approximated with a negative-binomial distribution compared to a Poisson distribution.

**Supplementary Figure 2.**
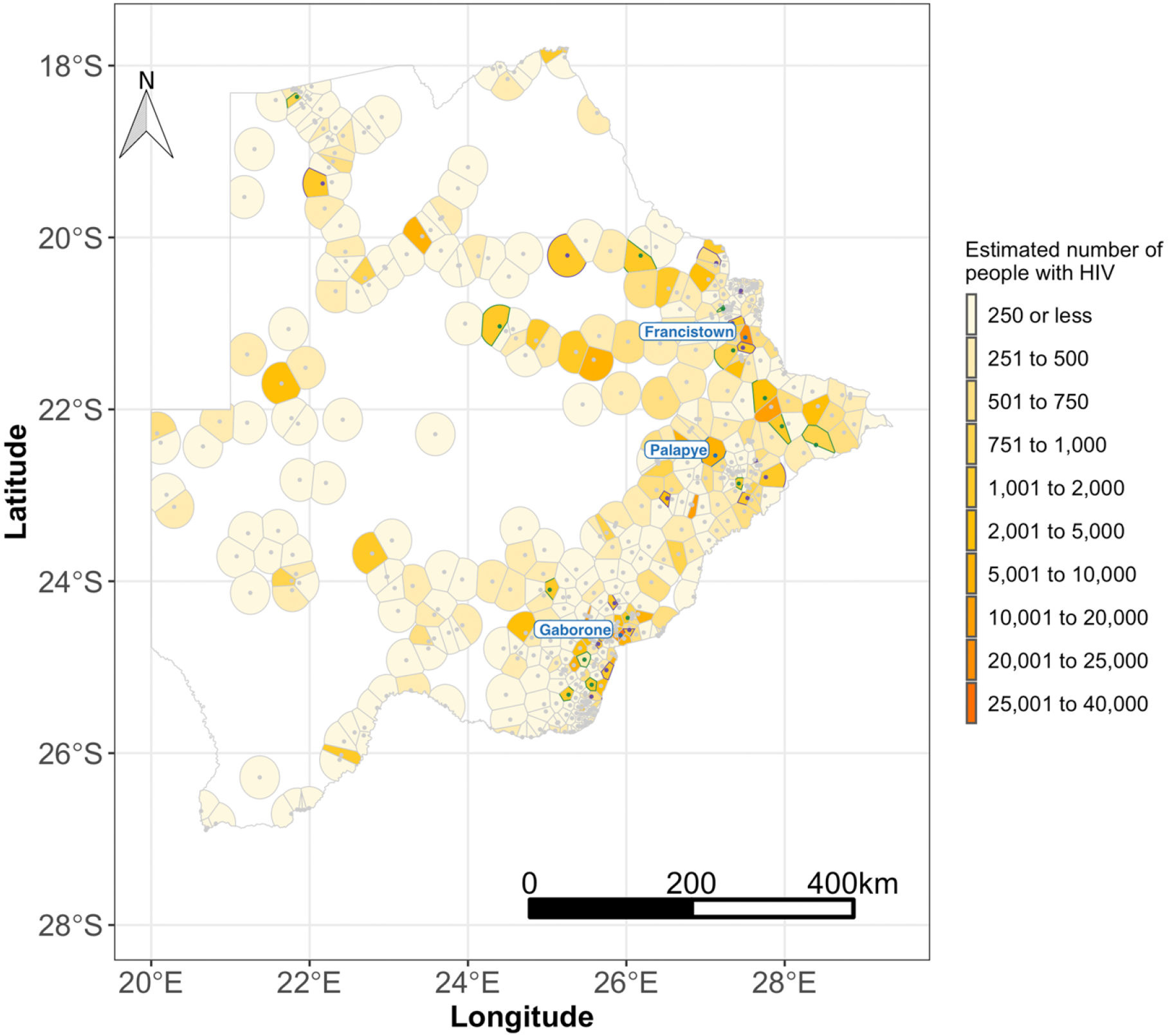
Spatial distribution of the estimated number of people with HIV-1 in Botswana. Estimates of the number of people with HIV-1 were computed from district HIV-1 prevalence estimates from the 2013 Botswana AIDS Impact Survey (BAIS 2013) and community-size estimates from the 2011 Botswana population and housing census. Intervention communities in the BCPP trial are denoted by purple filled circles and boundaries and control communities are represented by green filled circles and boundaries. The communities in the BCPP trial are distributed around three major urban areas: Gaborone city, Palapye and Francistown city represented by blue filled circles and labels.

**Supplementary Figure 3.**
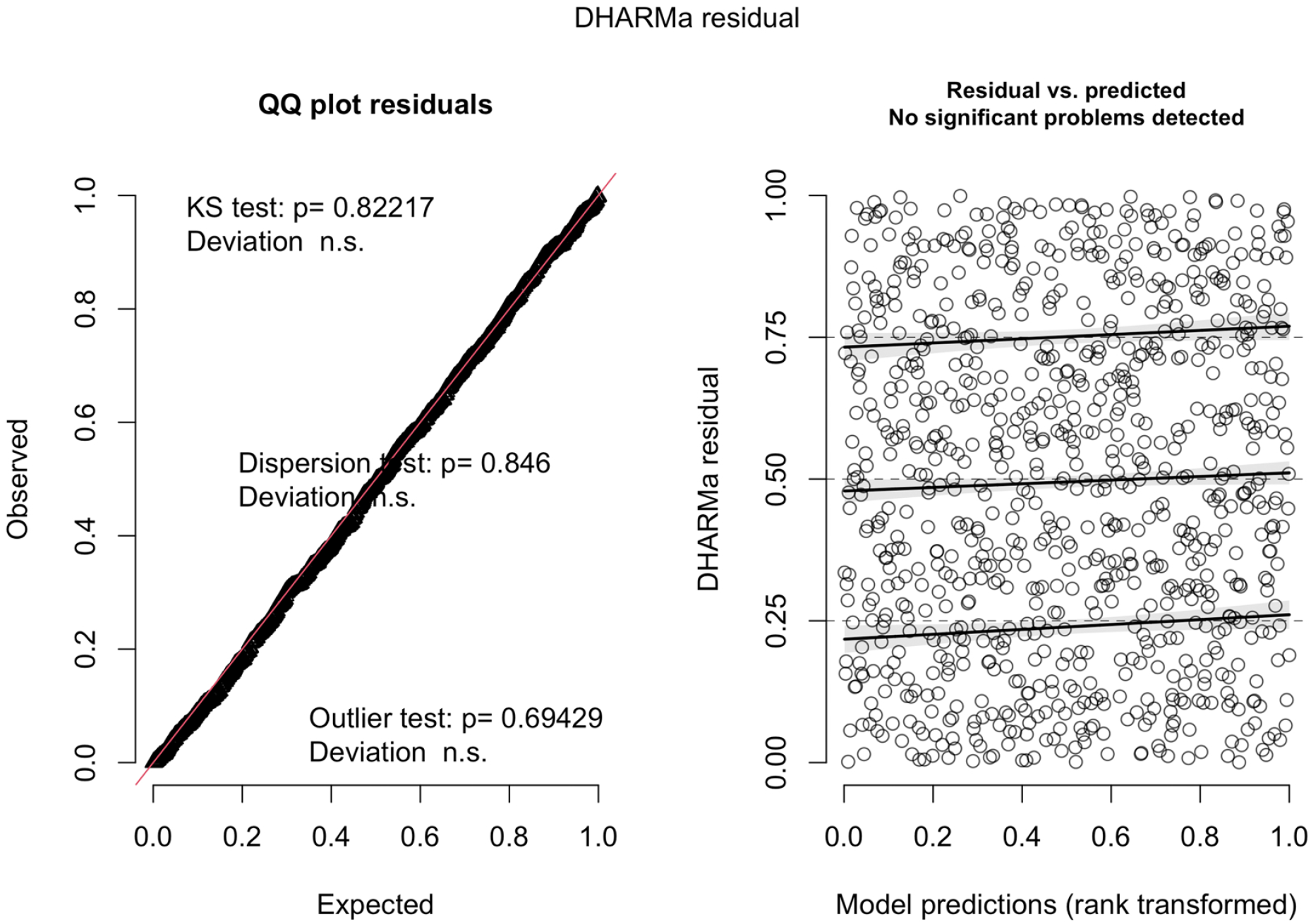
A quantile-quantile (QQ) residual plot that compares the distribution of residuals of 31 opposite-sex transmission pairs identified between ordered pairs of communities in the BCPP trial during the post-baseline period with those that would be expected under the post-baseline model in Table 1.

**Supplementary Figure 4.**
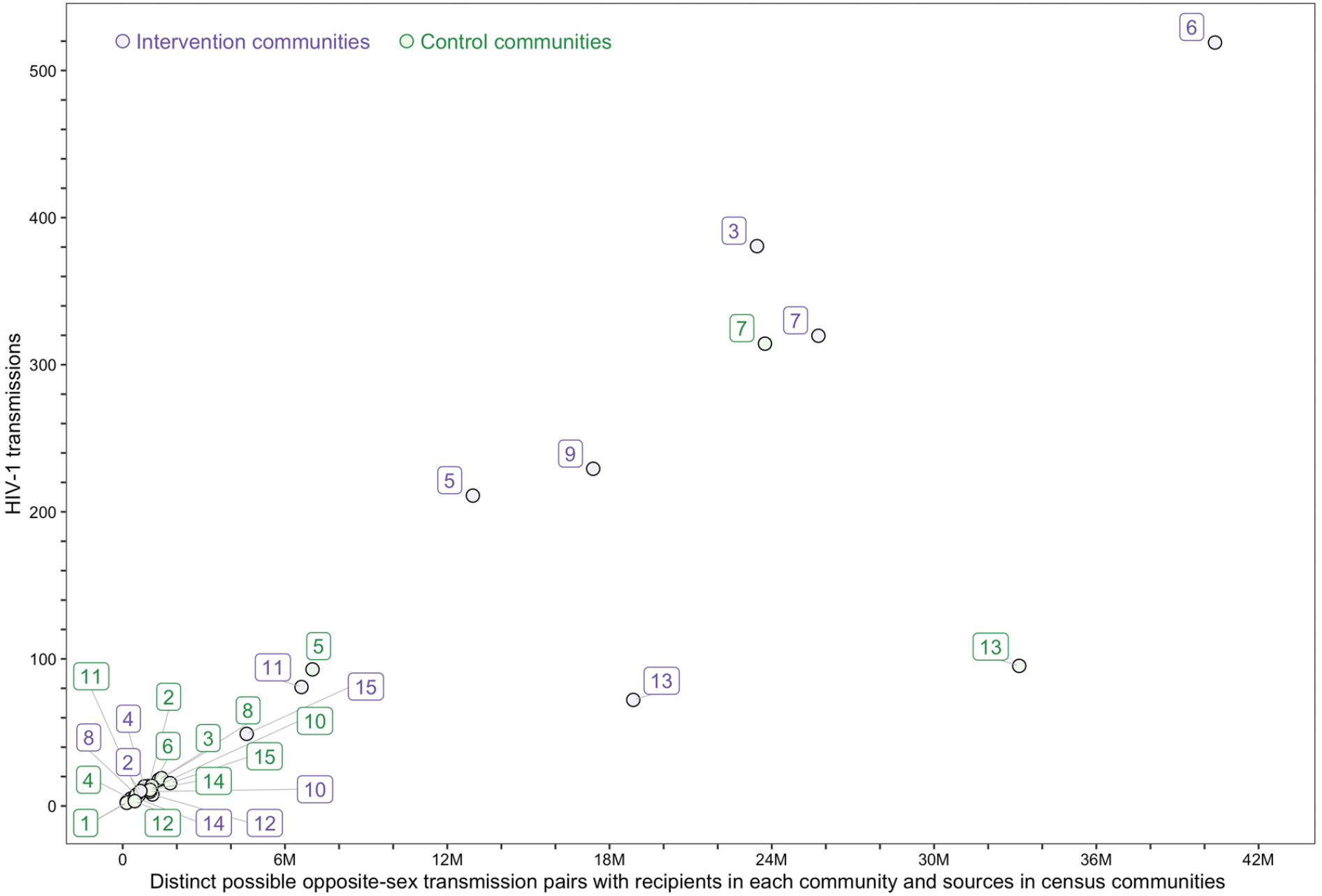
A scatter plot that shows a positive correlation between the estimated number of HIV-1 transmissions to recipients in trial communities that were predicted by the post-baseline model in Table 1 in the absence of the intervention and the total possible distinct opposite-sex transmission pairs that involve recipients in trial communities. The maximum (or total) distinct possible opposite-sex transmission pairs that involve recipients in trial communities represent the opportunity for transmission to recipients in the the trial communities, and are computed from the community size and HIV-1 prevalence of the source community and the number of people with HIV-1 sampled from the recipient community during the post-baseline period when the intervention could have taken effect. The BCPP trial matched communities into 15 pairs based on geographical proximity to major urban areas, population-size and age structure, and access to health services.

